# Impact of Viz LVO on Time-to-Treatment and Clinical Outcomes in Large Vessel Occlusion Stroke Patients Presenting to Primary Stroke Centers

**DOI:** 10.1101/2020.07.02.20143834

**Authors:** Jacob R. Morey, Emily Fiano, Kurt A. Yaeger, Xiangnan Zhang, Johanna T. Fifi

**Author notes:** **Corresponding Author:** Johanna T. Fifi, MD, Department of Neurosurgery, Icahn School of Medicine at Mount Sinai, The Mount Sinai Hospital, Klingenstein Clinical Center, 1-N, 1450 Madison Ave, New York, NY 10029.

## Abstract

**Introduction:** Randomized controlled trials have demonstrated the importance of time-to-treatment on clinical outcomes in large vessel occlusion (LVO) stroke. Delays in interventional radiology (INR) consultation are associated with a significant delay in overall time to endovascular treatment (EVT). Delays in EVT are particularly prevalent in Primary Stroke Centers (PSC), hospitals without thrombectomy capability onsite, where the patient requires transfer to a Thrombectomy Capable or Comprehensive Stroke Center for EVT. A novel computer aided triage system, Viz LVO, assists in early notification of the PSC stroke team and affiliated INR team. This platform includes an image viewer, communication system, and an artificial intelligence algorithm that automatically identifies suspected LVO strokes on CTA imaging and rapidly triggers alerts.

**Hypothesis:** Viz LVO will decrease time-to-treatment and improve clinical outcomes.

**Methods:** A prospectively maintained database was assessed for all patients who presented to a PSC currently utilizing Viz LVO in the Mount Sinai Health System in New York and underwent EVT following transfer for LVO stroke between October 1, 2018 and March 15, 2020. There were 42 patients who fit the inclusion criteria and divided into pre- and post-Viz ContaCT implementation by comparing the periods of October 1, 2018, to March 15, 2019, “Pre-Viz”, and October 1, 2019, to March 15, 2020, “Post-Viz.” Time intervals and clinical outcomes were collected and compared.

**Results:** The Pre- and Post-Viz cohorts were similar in terms of gender, age, proportion receiving IV-tPA, and proportion with revascularization of TICI > 2B. The presenting NIHSS and pre-stroke mRS scores were not statistically different.

The median initial door-to-INR notification was significantly faster in the post-Viz cohort (21.5 minutes vs 36 minutes; p=0.02). The median initial door-to-puncture time interval was 20 minutes shorter in the Post-Viz cohort, but this was not statistically significant (p=0.20).

The 5-day NIHSS and discharge mRS were both significantly lower in the Post-Viz cohort (p=0.02 and p=0.03, respectively). The median 90-day mRS scores were also significantly lower post-Viz implementation, although a similar proportion received a good outcome (mRS score ≤ 2) (p=0.02 and p=0.42, respectively).

**Conclusions:** EVT is a time-sensitive intervention that is only available at select stroke centers. Significant delays in time-to-treatment are present when patients require transfer from PSCs to a EVT capable stroke center. In a large health care system, we have shown that Viz LVO implementation is associated with improved time to INR notification and clinical outcomes. Viz LVO has the potential for wide-spread improvement in clinical outcomes with implementation across large hub and stroke systems across the country.

## Introduction

Numerous randomized controlled trials and meta-analyses have demonstrated the importance of time to treatment on patient outcomes in large vessel occlusion (LVO) stroke.^1^ Delays in neurointervention consultation are associated with a significant delay in overall time to endovascular treatment. Delays in endovascular treatment are particularly prevalent in Primary Stroke Centers (PSC), hospitals without thrombectomy capability onsite, where the patient requires transfer to a Thrombectomy Capable or Comprehensive Stroke Center for treatment.^2^ A novel Computer Aided Triage System, Viz LVO, offers a technological solution to enable early notification of the PSC stroke team and affiliated neurointerventional team, and is hypothesized to improve times to treatment and patient outcomes in patients with large vessel occlusions.

### Device Description

The Viz Neuroimaging Platform (Viz.ai®, Palo Alto, CA) was designed to optimize the assessment and workflow of acute ischemic stroke (AIS) patients with a special focus on potential candidates for acute reperfusion treatments. It received FDA clearance as Viz ContaCT. The system is based on a proprietary artificial intelligence (AI) convolutional neural network trained using machine learning techniques for automated neuroimaging processing and interpretation. The clinical application tool is constituted by a HIPAA-compliant mobile interactive module encompassing (1) a Digital Imaging and Communications in Medicine (DICOM) viewer, (2) a communication system including text messaging as well as multiuser audio and video calling capabilities that enables the secure communication of sensitive medical information and follows the clinical workflow, (3) an AI algorithm that automatically identifies suspected large vessel occlusion (LVO) strokes on CTA imaging (with an accuracy of approximately 90% in cases of proximal anterior circulation LVO) and rapidly triggers an alert to the on-call team.

The Viz LVO Platform was implemented at Mount Sinai Health System in New York in September 2019.

## Methodology

An institutional, prospectively maintained database was assessed for all patients who presented to a Primary Stroke Center currently utilizing Viz in the Mount Sinai Hospital Network in New York and subsequently underwent mechanical thrombectomy following transfer for LVO stroke between October 1, 2018 and March 15, 2020. Institutional review board approval was obtained for this study and the need for patient consent was waived. Data were compared pre-Viz ContaCT implementation and post-Viz ContaCT implementation by comparing the periods of October 1, 2018 to March 15, 2019, “Pre-Viz”, and October 1, 2019 to March 15, 2020, “Post-Viz.” Overall initial door-to-treatment time and patient outcomes were collected and compared.

All patients who presented with an initial diagnosis of acute stroke to a Mount Sinai PSC emergency department and were subsequently transferred for thrombectomy during this period were included. A total of 42 patients were analyzed, with a mean age of 73.9 years. Summary statistics included age, gender, IV-tPA given, median initial door-to-puncture time, median door-to-interventional neuroradiologist (INR) notification, revascularization, Thrombolysis in cerebral infarction (TICI) score, and patient outcomes, as measured by 5-day NIHSS and discharge and 90-day mRS. Values were compared pre-Viz and post-Viz implementation.

For binary variables, Fisher’s exact test was performed. For continuous variables, the Mann-Whitney *U* test and t-test were performed. The data was reported as counts with percentages for categorical values and mean +/- standard deviation and median for continuous variables where appropriate. A value of p<0.05 was noted as statistically significant. Analyses were performed using SAS, Version 9.4.

## Results

The Pre- and Post-Viz cohorts were similar in terms of gender, age, percentage receiving IV-tPA, and percentage with revascularization of TICI > 2b. The initial presenting NIHSS and pre-stroke mRS scores were not statistically different between both cohorts (Table).

**Table:** Baseline Characteristics, Time Intervals, and Clinical Outcomes

| Variable | Pre-Viz | Post-Viz | P Value |
| --- | --- | --- | --- |
| **Baseline Characteristics** |  |  |  |
| Gender (female) | 45.0 (9/20) | 50.0 (11/22) | 0.7675 |
| Age | 75.8 +/- 15.35 | 72.1 +/-16.51 | 0.4566 |
| IV-tPA (%) | 30.0 (6/20) | 33.3 (7/21) | 1.00 |
| Revascularization (TICI > 2b) | 65.0 (13/20) | 55.0 (11/20) | 0.7475 |
| Initial NIHSS | 17.5 +/- 5.84 | 13.16 +/- 7.54 | 0.0591 |
| Pre-stroke mRS | 1.25 +/- 1.52 | 0.68 +/- 0.99 | 0.1554 |
| **Time Intervals** |  |  |  |
| Initial Door-to-Puncture (Median) | 185 | 165 | 0.2032 |
| Initial Door-to-Puncture (Mean+/-std) | 197.35 +/- 72.02 | 163.59 +/- 32.74 | 0.0655 |
| Initial Door-to-TICI 2B+(Mean+/-std) | 227.3 +/-78.80 | 206.8 +/- 57.54 | 0.3562 |
| Initial Door-to-TICI 2B+(Median) | 409.0 | 336.0 | 0.4396 |
| Initial Door-to-INR Notification (Median) | 36 | 21.5 | **0.0233** |
| Initial Door-to-INR Notification (Mean+/-std) | 55.2 +/- 47.69 | 24.86 +/- 12.42 | **0.0116** |
| CTA-to-Puncture (Median) | 154 | 143.5 | 0.4070 |
| CTA-to-Puncture (Mean+/-std) | 161+/-48.5 | 147+/-32 | 0.2869 |
| Door-In Door-Out (Median) | 106 | 94 | 0.1031 |
| Door-In Door-Out (Mean+/-std) | 122.75 +/- 60.5 | 94.16 +/- 33.96 | 0.0769 |
| **Clinical Outcomes** |  |  |  |
| 5-Day NIHSS | 21.93 +/- 14.66 | 10.78 +/- 12.25 | **0.0235** |
| Discharge mRS | 4.62 +/- 1.61 | 2.92 +/- 2.14 | **0.0318** |
| 90-Day mRS (Mean) | 4.74 +/- 1.73 | 3.40 +/- 1.96 | **0.0424** |
| 90-Day mRS (Median) | 5 | 3 | **0.0219** |
| mRS score ≤ 2 at 90 days | 15.79 (3/19) | 33.33 (5/15) | 0.4172 |

The median initial door-to-INR notification was significantly faster in the post-Viz cohort (21.5 minutes vs 36 minutes; p = 0.02) (Table). The median Initial door-to-puncture time interval was 20 minutes shorter in the Post-Viz cohort, as compared to the Pre-Viz cohort, but this was not considered significant (p = 0.20).

The 5-day NIHSS and discharge mRS were both significantly lower in the Post-Viz cohort, as compared to the Pre-Viz cohort (p = 0.02 and 0.03, respectively). The long-term median 90-day mRS scores were also significantly lower post-Viz implementation (p = 0.02). The proportion who received a good outcome (mRS score ≤ 2), was higher post-Viz, but the difference was not significant (p = 0.42) (Table).

## Discussion

In LVO stroke patients presenting to a Primary Stroke Center and subsequently transferred to a CSC or TSC for thrombectomy, there was a significant reduction in initial door-to-neurointerventionalist notification (p = 0.02) and a significant improvement in long-term outcomes following the implementation of Viz LVO (p = 0.02). We believe these clinical outcome improvements result from the early notification of the stroke and neuroendovascular teams, allowing early involvement in the care of LVO stroke patients, and therefore faster time to treatment.

The trends in decreased time-to-treatment leading to improved clinical outcomes in this study are similar to previous studies.^2-4^ The current study focused on transfer patients, which presents a great opportunity to decrease time-to-treatment, but its application may be used for evolving stroke systems of care using Mobile Interventional Stroke Teams traveling to thrombectomy capable stroke centers, as well as Mothership models.^5,6^

### Limitations

The major limitations of this study are the observational design and small, preliminary sample size.

## Conclusions

Endovascular therapy is a time-sensitive intervention that is only available at select stroke centers. Significant delays in time-to-treatment are present when patients require transfer from Primary Stroke Centers to those with endovascular capabilities. In a large health care system, we have shown that Viz LVO implementation is associated with improved time to treatment and clinical outcomes. Viz LVO allows for rapid viewing of imaging, coordination of teams, and an AI algorithm that automatically identifies and alerts teams to suspected LVO strokes on CTA imaging. It represents a novel application of artificial intelligence that acts as an early warning system and serves as a fail-safe to ensure that the INR team is notified earlier and LVO stroke patients are not missed or receive delayed treatment. This has the potential for wide-spread improvement in clinical outcomes with implementation across large hub and stroke systems across the country.

## Data Availability

The data that support the findings of this study are available from the corresponding author upon reasonable request.

## Notes

### Competing Interest Statement

J.T. Fifi: Research Grant; Significant; Stryker. Other - Consultant for research trial; Modest; Stryker. Other - Faculty at Fellow's Course; Significant; Microvention. Other - Consultant for research trial; Modest; Penumbra. Stock Shareholder; Significant; Synchron, Cerebrotech, Endostream, Imperative Care. Research Grant; Significant; Penumbra.

### Funding Statement

No external funding received.

### Author Declarations

Institutional review board approval was obtained for analysis of the dataset.

